# Health system and household costs of preventive treatment delivery for children exposed to multidrug-resistant tuberculosis in South Africa

**DOI:** 10.1101/2025.10.29.25339111

**Authors:** Thomas Wilkinson, Suereta Fortuin, Anneke C. Hesseling, H. Simon Schaaf, Trinh Duong, Diana M. Gibb, Susan E Purchase, Lee Fairlie, Suzanne Staples, Neil Martinson, Edina Sinanovic, James A. Seddon

**Affiliations:** School of Public Health and Family Medicine, University of Cape Town, Health Economics Unit, Cape Town, South Africa; Division Molecular Biology and Human Genetics, Stellenbosch University, Cape Town, South Africa; Desmond Tutu TB Centre, Stellenbosch University, Cape Town, South Africa; Institute of Clinical Trials and Methodology, Faculty of Population Health Sciences, University College London, United Kingdom; MRC Clinical Trials Unit, University College London, London, United Kingdom; Wits RHI, University of Witwatersrand, Johannesburg, South Africa; THINK, THINK South Africa, Durban, South Africa; Perinatal HIV Research Unit (PHRU) and Infectious Diseases and Oncology Research Institute, University of Witwatersrand, Johannesburg, South Africa; Division of Infectious Diseases, School of Medicine, Johns Hopkins University, Baltimore, USA; Imperial College London, Department of Infectious Disease, London, United Kingdom of Great Britain and Northern Ireland

**Keywords:** Costing, Pediatric, Children, Multidrug-resistant tuberculosis, Tuberculosis preventive therapy

## Abstract

**Background:** Tuberculosis (TB) preventive treatment (TPT) is a core component of TB control. Levofloxacin was recently recommended by the World Health Organization as TPT in children exposed to multidrug-resistant tuberculosis (MDR-TB). However, the affordability and feasibility of TPT provision for governments, health services, individual households and global donors is uncertain.

**Methods:** TB-CHAMP was a cluster-randomized, double-blind, placebo-controlled trial investigating levofloxacin TPT in children exposed to MDR-TB in South Africa and included cost analysis of TPT delivery. We modelled the societal (health system and household) cost of levofloxacin TPT, other potential future (but not currently recommended) MDR-TB exposure TPT options, and compared with drug-susceptible TPT options in children and adolescents under 15 years old. We modelled costs across 12 different medicine and formulation options for 11 paediatric dosing weight-bands using TB-CHAMP and household survey data, with national and international procurement medicine pricing.

**Results:** Total societal costs of providing levofloxacin TPT and its monitoring per child were US$252 (0-<5-years) and US$256 (5-<15-years) and had comparable costs to drug-susceptible TPT regimens when using Global Drug Facility pricing. Pharmaceutical costs for a course of TPT using paediatric formulations in children <5 years varied between US$38 (levofloxacin), US$80 (bedaquiline) and US$582 (delamanid). Household costs associated with accessing TPT and ongoing monitoring ranged from 1.6%-2.1% of median annual income of affected households, with shorter TPT courses incurring lower household costs. Global Drug Facility procurement achieved TPT pricing at 70% of the costs of national South African procurement, and enabled access to a wider range of child-friendly formulations.

**Discussion:** Costs of TPT are influenced by factors including medicine pricing and procurement, the use of child-friendly formulations, implementation approach, the duration of treatment, and choice of TPT regimen across children’s weight and age distributions. Costs to households are relatively small but may be significant for impoverished households already managing the financial disruption of a family member with MDR-TB disease

## INTRODUCTION

Each year, an estimated 1.3 million children under 15 years of age develop tuberculosis (TB) worldwide, causing 240,000 deaths, 80% in children under five (1). Preventing TB in at-risk children is a core pillar of the global TB response (2), and includes identification of child household contacts of people with TB disease, screening for active TB disease, and providing TB preventive treatment (TPT) where appropriate. TPT involves providing a course of antituberculosis antibiotics to lower the risk of developing TB disease; however, achieving universal coverage for children remains a major implementation challenge. Between 2018 and 2022, only 40% of the targeted four million eligible children globally initiated TPT (3).

While TPT is widely recommended for contacts of patients with drug-susceptible TB, the optimal approach for child contacts of people with multidrug-resistant (MDR)-TB —defined as *Mycobacterium tuberculosis* resistant to at least isoniazid and rifampicin—has been less certain given the lack of robust data. Evidence has now emerged from recent trials. TB-CHAMP, a phase III trial in South Africa, evaluated six months of daily levofloxacin versus placebo in children (4). V-QUIN, conducted in Viet Nam, assessed levofloxacin for 24 weeks in all age groups (5). The PHOENIx trial is underway in 12 countries, comparing delamanid with isoniazid TPT in all high-risk household contacts, including children (6). Additional agents are under investigation including the use of bedaquiline (7) (8), and observational studies have demonstrated isoniazid efficacy in child contacts of MDR-TB index patients (9) (10).The findings from TB-CHAMP and V-QUIN have informed the 2024 World Health Organization (WHO) recommendation for a six-month levofloxacin regimen for household contacts of people with MDR-TB (11). In addition to establishing the most effective anti-TB agents for children, emerging evidence is demonstrating the importance of the type of formulation, palatability, and regimen length as key elements of effective and feasible TB treatment and prevention for children (12) (13).

Introducing MDR-TB TPT in children has important cost implications. Countries transitioning from donor support face particularly difficult trade-offs, as health investment compete for constrained budgets. Decision makers are also concerned about household costs: although TB services are free from direct cost at the point of care in South Africa and many other settings, accessing treatment can impose a substantial financial burden, especially in households already managing the costs of a family member’s MDR-TB treatment (14). Although eliminating catastrophic TB-related costs is a main target of the WHO End TB Strategy (15), there is limited evidence on the costs to households associated with a child accessing MDR-TB treatment. A household survey, MDRTBkids, was conducted in South Africa in 2021. Despite free TB treatment at the point of use in South Africa, the study found that the median direct and indirect cost to households for accessing MDRTB treatment for a child was more than US$500 per episode of care, and resulted in more than half of households experiencing catastrophic health expenditure, defined as more than 20% of annual household income (16).

Robust cost estimates are essential for planning and prioritising TPT implementation at national and global level. Although global systematic reviews (17) and standardised cost repositories (18) have improved access to TB cost data, there remains little localised information on the costs of providing MDR-TB TPT for children and adults. In addition to health system costs, pharmaceutical pricing and formulation availability vary between countries, shaped by national procurement mechanisms. The Global Drug Facility (GDF) offers pooled procurement to secure affordable, quality-assured TB medicines (19). This is particularly relevant for child-friendly TB medicines, where the market size in any one country may limit ability to achieve reasonable prices or a range of appropriate formulations. Countries such as South Africa, which procure through national tenders, do not routinely access GDF prices or formulations, which impacts on the expected costs of TPT implementation and range of formulations available for children in different age and weight ranges.

## METHODS

This study assesses the cost of delivering TPT for children <15 years exposed to MDR-TB from a health system and societal perspective in the South African context, reflecting differences in costs associated with weight-based dosing, formulations, and procurement mechanisms. The study incorporates empirical data from the TB-CHAMP trial, the MDRTBkids household survey, normative South African and WHO guidelines, medicine pricing from South Africa’s national procurement programme and the GDF, and South Africa-specific health system costs. See Table 1 for the parameter inputs used in the analysis.

**Table 1.** Parameter cost inputs.

|  | Value |  | Reference |
| --- | --- | --- | --- |
| **MEDICINE COSTS** | *Global Drug Facility* | *South African national procurement* |  |
| Levofloxacin 500mg tablet | US\$ 0.05 | US\$0.13 | (28) (27) |
| Levofloxacin 250mg tablet | US\$ 0.03 | US\$ 0.10 | (28) (27) |
| Levofloxacin 100mg dispersible tablet | US\$ 0.12 | - | (28) |
| Isoniazid 100mg tablet | US\$ 0.01 | US\$ 0.03 | (28) (27) |
| Rifampicin 75mg/Isoniazid 50mg FDC tablet | US\$ 0.05 | US\$ 0.05 | (28) (27) |
| Rifampicin 150mg/Isoniazid 75mg FDC tablet | US\$ 0.05 | US\$ 0.06 | (28) (27) |
| Rifampicin 150mg capsule | US\$ 0.14 | US\$ 0.10 | (28) (27) |
| Rifapentine 150mg tablet | US\$ 0.30 | US\$ 0.30 | (28) (27) |
| Delamanid 50mg tablet | US\$ 1.48 | US\$ 1.69 | (28) (27) |
| Delamanid 25mg dispersible tablet | US\$ 1.77 | - | (28) |
| Bedaquiline 100mg tablet | US\$ 0.38 | US\$ 0.61 | (28) (27) |
| Bedaquiline 20mg tablet | US\$ 0.43 | - | (28) |
| **HEALTH SERVICES COSTS** |  |  |  |
| Standard PHC clinic visit | US\$ 12.47 | | (36) (31) |
| Medicine dispensing fee | US\$ 2.60 | | (30) |
| Chest X-ray | US\$ 66.64 | | (32) |
| HIV test | US\$ 2.86 | | (29) |
| Primary Health Care clinic screening visit:<br>*Clinic visit + Chest X-ray + HIV test + dispensing fee.* | US\$ 84.51 | | Calculated |
| Primary Health Care clinic TPT visit:<br>*Clinic visit + dispensing fee* | US\$ 15.07 | | Calculated |
| **HOUSEHOLD COSTS AND INPUTS** |  |  |  |
| Average travel cost to visit clinic (round trip) | US\$ 0.59 | | (16) |
| Average costs incurred at the clinic (food) | US\$ 1.01 | | (16) |
| Average travel time to visit clinic (round trip) | 40 minutes |  | (16) |
| Average time spent at clinic per visit | 97 minutes |  | (16) |
| Minimum wage – South Africa 2025 (per hour) | US\$ 1.56 | | (37) |
| Time cost of one PHC visit | US\$ 3.55 | | (16) |
| **EPIDEMIOLOGY AND RESOURCE UTILISATION** |  |  |  |
| Proportion that will require HIV testing | 97.9% |  | (4) |
| Proportion of dispensing clinic visits attended | 97.8% |  | (4) |

PHC = Primary Health Care; FDC = Fixed dosed combination (tablet)

A patient care pathway for household contacts of people diagnosed with MDR-TB in South Africa was developed using local guidance (20), informed by the TB-CHAMP trial protocol, but aimed to reflect a routine programmatic implementation scenario and excluded trial-specific elements such as additional blood tests. The patient pathway incorporates screening for TB, then either, referral for initiation of TB treatment (in case of disease) or offering TPT, and an observation period (Figure 1).

**Figure 1.**
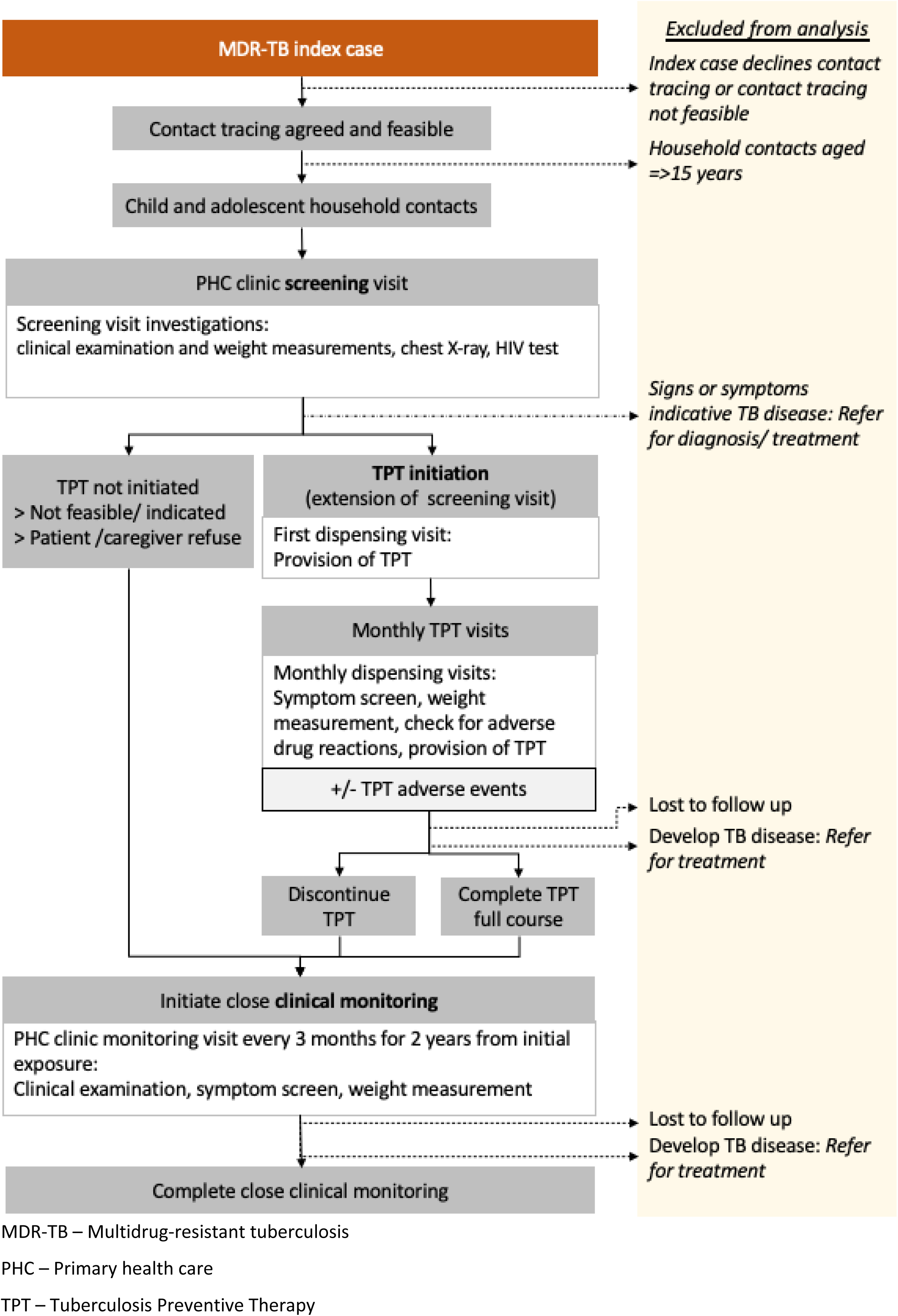
Care pathway of MDR-TB contacts.

An ingredients-based approach was used to incorporate health system and household costs along the patient pathway (Supplementary appendix p.5). The TB-CHAMP trial indicated that levofloxacin was safe in children and demonstrated no statistically significant difference between levofloxacin and placebo at all grades of adverse events (4). The TB CHAMP trial also demonstrated that the adult formulation of the levofloxacin formulation was well-tolerated by children (21,22). The cost implications of high-grade adverse events commonly include additional clinic or hospital visits, investigations and treatment where required. The analysis assumed that the adverse event rate for all TPT options were as observed in TB-CHAMP trial, which included 89% children below 5 years of age. As further evidence emerges for the adverse effect profile of levofloxacin and other TPT options in children, any costs associated with adverse effect management, particularly in older age groups that may be of increased risk of adverse events, would need to be incorporated in future analysis.

### Healthcare costs approaches/sources

The analysis represented 12 different regimen and formulation combinations to reflect the range of current and future potential TPT options available in the 0 - <15-year age group (see Table 2). Total per-patient costs for each TPT option were calculated using a stepwise process by cost category.

**Table 2.** Overview of tuberculosis exposure management options.

| TB exposure management options | TPT formulation provided | Child-friendly formulation | TPT dosing period (months) | Clinical monitoring period (months) | TPT visits (n) | Clinical monitoring visits (n) |
| --- | --- | --- | --- | --- | --- | --- |
| Clinical monitoring only | None | N/A | N/A | 24 | 0 | 8 |
| **MDR/RR-TB exposure: (current recommendation)** |  |  |  |  |  |  |
| 6LFX_500mg tab | Levofloxacin 500mg tablet | No | 6 | 18 | 5 | 4 |
| 6LFX_250mg tab | Levofloxacin 250mg tablet | No |  |  |  |  |
| 6LFX_100mg disp tab | Levofloxacin 100mg dispersible tablet | Yes |  |  |  |  |
| 6LFX_optimised | Levofloxacin 100mg dispersible tablet (<25 kg); Levofloxacin 500mg tablet (25kg and over); | Partly (100mg dispersible tab) |  |  |  |  |
| 6hH_100mg tab<sup>1</sup> | Isoniazid 100mg tablet | Yes | 6 | 18 | 5 | 4 |
| **MDR/RR-TB exposure: (Potential future option)** |  |  |  |  |  |  |
| 6DLM_50mg tab | Delamanid 50mg tablet | No | 6 | 18 | 5 | 4 |
| 6DLM_25mg disp tab | Delamanid 25mg dispersible tablet | Yes |  |  |  |  |
| 6DLM_optimised | Delamanid 25mg dispersible tablet (<25 kg); Delamanid 50mg tablet (25kg and over); | Partly (25mg dispersible tab) |  |  |  |  |
| 6BDQ_100mg tab | Bedaquiline 100mg tablet | No | 6 | 18 | 5 | 4 |
| 6BDQ_20mg tab | Bedaquiline 20mg tablet | Yes |  |  |  |  |
| 6BDQ_optimised | Bedaquiline 20mg tablet (<25 kg); Bedaquiline 100mg tablet (25kg and over) | Partly (20mg dispersible tab) |  |  |  |  |
| **DS-TB exposure:** |  |  |  |  |  |  |
| 6stdH_100mg tab<sup>1</sup> | Isoniazid 100mg tablet | Yes | 6 | 18 | 5 | 4 |
| 4R_150mg cap | Rifampicin 150mg capsule | No | 4 | 20 | 3 | 5 |
| 3HR_50/75mg FDC tablet or 3HR_75/150 mg FDC tablet | Isoniazid 50mg / rifampicin 75mg FDC tablet (<25kg); Isoniazid 75mg / rifampicin 150mg FDC tablet (25kg and over) | Yes | 3 | 21 | 2 | 5 |
| 3HP_100+150mg tab<sup>2</sup> | Isoniazid 100mg tablet plus rifapentine 150mg tablet | No | 3 | 21 | 2 | 5 |

BDQ-bedaquiline, DLM-delamanid, FDC-fixed dose combination, hH-high dose isoniazid, stdH-standard dose isoniazid, LFX-levofloxacin, P-rifapentine, R-rifampicin; TPT-tuberculosis preventive therapy. All options receive 1 screening visit.
1. 9H (9 months of isoniazid daily) and 12H (12 months of isoniazid daily) are both recommended TPT options, but 6H (6 months of isoniazid daily) was selected for this analysis as it most closely reflects current clinical practice in South Africa. 2. 3HP is not a recommended TPT option for children weighing <25kg in the 'National Guideline on the treatment of tuberculosis infection in South Africa; but was included in this analysis as it is a WHO-recommended TPT option for drug-susceptible tuberculosis in children and the South African guideline notes recommendations may change once more safety data on 3HP becomes available.

Numbers of tablets or capsules were calculated according to the weight-based harmonised dosing recommended in the WHO consolidated guidelines on TPT for standard-dose isoniazid, rifapentine, rifampicin, levofloxacin (23) and the South African guidelines for delamanid and high-dose isoniazid (20). Recommended therapeutic treatment doses were used to reflect TPT dosing for bedaquiline (25). We assumed medicines were dispensed for 28 days at a time, and that the required dose administered was rounded up to the nearest whole tablet, or half tablet where tablets are scored or are dispersible (26). As older children may be more able to tolerate a solid dosage tablet formulation, we modelled an “optimised” regimen using the dispersible tablet in the lower weight bands (<25 kg), and solid dosage in the higher weight bands. The rate of scheduled medicine dispensing and monitoring visits attended by participants in the TB-CHAMP trial were used to reflect cost implications expected during implementation. The reported medication adherence (defined as proportion of prescribed tablets consumed) in the TB-CHAMP trial was high (96%; interquartile range 90%-99%) with 73% of participants taking more than 90% of prescribed doses, and no significant difference between adherence for levofloxacin compared to placebo (4). The rate of attendance to medicine-dispensing visits in TB-CHAMP (97.8%) was applied to the calculated cost of all TPT regimens in this analysis to reflect the expected under-use of medicine in an implementation scenario, recognizing that once a monthly prescription is dispensed, the total of the cost of the medicine is incurred (under an assumption that a medicine that is dispensed but not consumed is wasted), but non-dispensed medicine does not incur a cost.

Unit costs of medicines were calculated using prices listed in South Africa’s public sector national procurement system October 2025 (27) and the GDF September 2025 (28), costs of laboratory tests were from the National Health Laboratory Service (NHLS) state pricing list (current version published in 2023 and are current in year 2025) (29), dispensing costs were sourced from the Uniform Patient Fee Schedule (UPFS) (April 2025) (30) and clinic visit and chest x-ray unit costs were derived from existing literature (31) (32). Expected frequency of routine clinic visits were calculated based on South Africa’s national TB guidance (20).

Non-medicine health system costs were determined by the type and number of contacts with the health service for each TPT option. All options included a screening visit, which consisted of a standard clinic visit (for symptom screening, weight measurement, physical examination), plus a chest x-ray, HIV test (if status unknown), and dispensing of the first month of TPT (except for the non-TPT option). Under TB CHAMP, a test of TB infection was not required in children < 5 years of age and in older children (with and without HIV) a test of TB infection was required. Current TB programme resource constraints mean that tests of TB infection are not routinely provided in South Africa (and therefore costs are not incurred) in those 5 years of age and older who are exposed to TB. Therefore, the base case costs represented in this analysis do not incorporate TST testing for any age groups and may be appropriate for estimating and implementation budget impact analysis. The costs of introducing TST for children 5 years old and above is provided as a sensitivity analysis and should be considered for cost effectiveness analysis using effect sizes as demonstrated in the TB CHAMP trial. Monthly TPT dispensing visits consisted of a standard clinic visit, checking for adverse drug reactions, weight gain and dispensing of TPT medicines and adjusting doses as required. Clinical monitoring visits consisted of a standard clinic visit and were estimated to take place every three months up to two years from initial exposure once the TPT course was completed, or monthly for six months followed by every three months for two years, if no TPT was provided. Healthcare costs associated with TPT provision excluded procedures and interventions required for the identification and management of TB disease and/or those required for the management and monitoring of HIV.

Costs collected from published literature were converted to South African Rand (ZAR) in the year of analysis. When relevant, costs were inflated to 2025 using the South African consumer price index (33). Results were developed in ZAR and US$ and are presented in the main text for international audience in US$ without adjustment for purchasing power parity.

The TB-CHAMP trial – the protocol of which included this analysis, was approved by the Stellenbosch University Health Research Ethics Committee (M16/02/009) and the University of the Witwatersrand Ethics Committee (160409) and the South African Health Products Regulatory Authority (SAHPRA). The MDRTBkids study received ethics approval from the Stellenbosch University Health Research Ethics committee (N20/09/102).

### Societal cost approach/sources

The household costs analysis utilises data collected from the MDRTBkids survey, a concurrent costing survey that included data from 45 households with a child with MDR-TB disease in the Western Cape, South Africa, who routinely initiated treatment between 1 January 2018 to 31 August 2021 (16). The costs incurred by households to access TPT and monitoring included travel costs, expenses while visiting facilities and time costs of attending clinics, and these were uplifted to 2025 costs by applying the annual South African consumer price index (33). The analysis estimated household costs by applying average per-instance cost of attending clinic visits and collecting prescribed medicines as observed in the MDRTBkids survey, multiplied by the expected health system interaction of the different TPT management strategies. While the number of dispensing and clinic visits varied between some strategies, it was assumed that per-clinic visit costs did not differ between TPT regimens – the same monthly collection cost would be incurred by households regardless of whether it was to collect a prescription for levofloxacin or for delamanid. The total average household costs of accessing TPT over the treatment and monitoring duration was represented as a proportion of average household income of participants in the MDRTBkids survey population, under an assumption that average monthly income of a household with a child with MDR-TB disease would be comparable to a household with a child who is exposed to MDR-TB.

## RESULTS

The pharmaceutical costs per patient by TPT regimen, formulation, age range, and procurement mechanism are shown in Table 3, the corresponding costs per weight band is shown in Table S8 supplementary appendix. Figure 2 shows levofloxacin 6-month TPT regimen cost by weight, with weighted average across the 0-<15-years age range.

**Figure 2.**
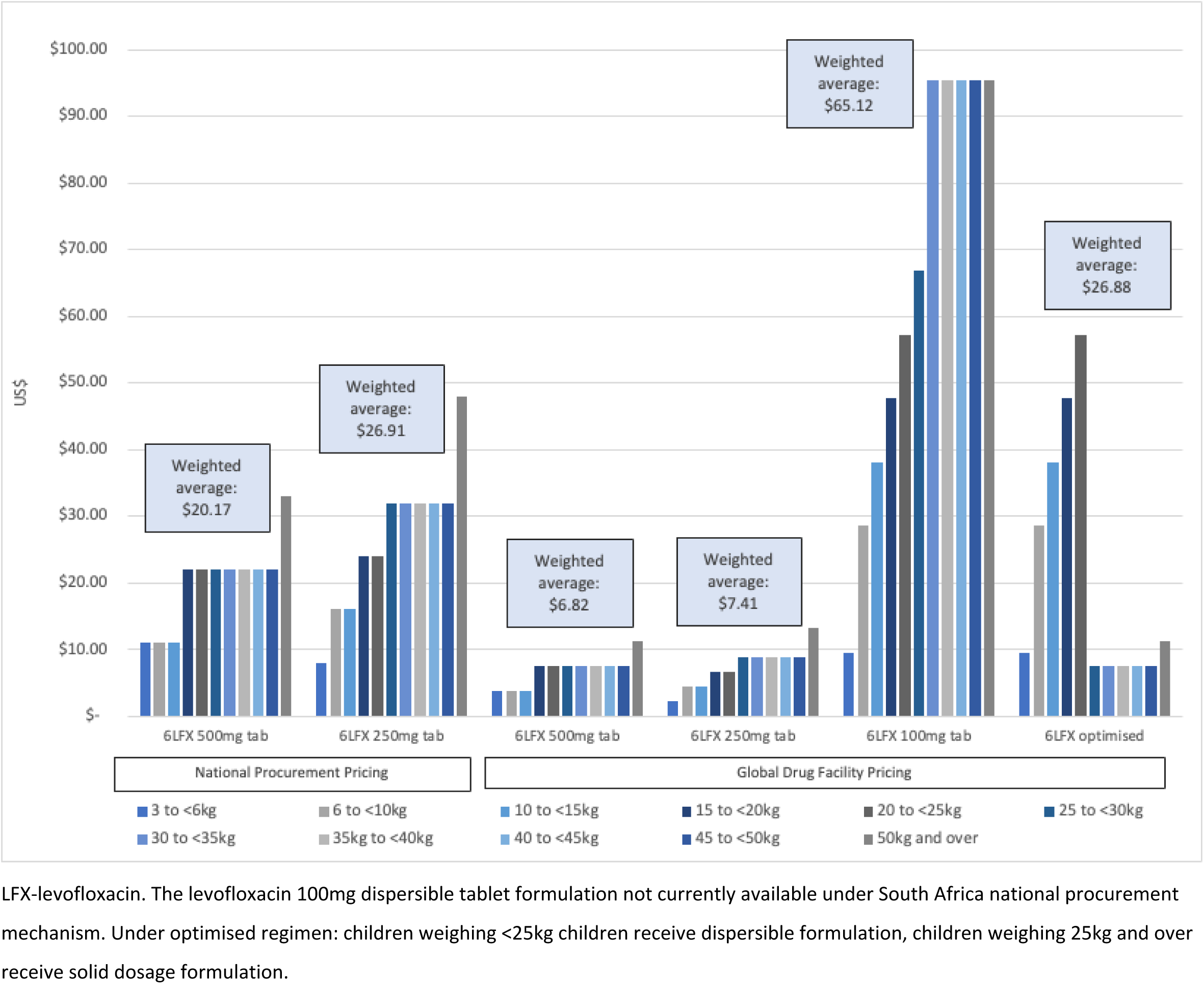
Medicine cost by weight band for 6-month levofloxacin-containing TPT options, with weighted average across the 0-<15-year-old age range.

**Table 3.** Pharmaceutical cost of TPT per patient by regimen, procurement mechanism and age cohort.

| | GDF price (\$) | | | | National price (\$) | | | |
| --- | --- | --- | --- | --- | --- | --- | --- | --- |
| TPT regimen | Age 0-<2 | Age 2-<5 | Age 0-<5 | Age 5-<15 | Age 0-<2 | Age 2-<5 | Age 0-<5 | Age 5-<15 |
| 6LFX_500mg tab | \$3.72 | \$5.31 | \$4.67 | \$7.84 | \$11.00 | \$15.70 | \$13.82 | \$23.18 |
| 6LFX_250mg tab | \$4.36 | \$5.35 | \$4.95 | \$8.65 | \$15.81 | \$19.41 | \$17.97 | \$31.41 |
| 6LFX_100mg disp tab | \$31.98 | \$42.45 | \$38.26 | \$79.03 | - | - | - | - |
| 6LFX_optimised | \$31.98 | \$42.33 | \$38.19 | \$21.27 | \$31.98 | \$42.36 | \$38.20 | \$32.47 |
| 6DLM_50mg tab | \$243.22 | \$243.22 | \$243.22 | \$552.65 | \$278.24 | \$278.24 | \$278.24 | \$632.21 |
| 6DLM_25mg disp tab | \$581.79 | \$581.79 | \$581.79 | \$1,321.93 | - | - | - | - |
| 6DLM_optimised | \$581.79 | \$581.12 | \$581.39 | \$648.86 | \$581.79 | \$581.19 | \$581.43 | \$718.48 |
| 6BDQ_100mg tab | \$29.67 | \$31.89 | \$31.01 | \$45.59 | \$47.99 | \$51.58 | \$50.14 | \$73.73 |
| 6BDQ_20mg tab | \$57.64 | \$94.69 | \$79.85 | \$233.45 | - | - | - | - |
| 6BDQ_optimised | \$57.64 | \$94.37 | \$79.66 | \$69.02 | \$57.64 | \$94.42 | \$79.69 | \$91.04 |
| 6hH_100mg tab | \$2.84 | \$4.10 | \$3.59 | \$7.17 | \$6.96 | \$10.05 | \$8.81 | \$17.57 |
| 6stdH_100mg tab | \$2.01 | \$2.89 | \$2.54 | \$4.31 | \$4.93 | \$7.08 | \$6.22 | \$10.57 |
| 4R_150mg cap | \$15.55 | \$16.08 | \$15.87 | \$44.93 | \$10.92 | \$11.29 | \$11.14 | \$31.56 |
| 3HR_FDC tab | \$10.64 | \$15.28 | \$13.42 | \$15.07 | \$10.53 | \$15.12 | \$13.28 | \$16.53 |
| 3HP_100mg + 150mg tab | \$6.13 | \$8.92 | \$7.80 | \$17.80 | \$6.52 | \$9.48 | \$8.30 | \$18.99 |

BDQ-bedaquiline, DLM-delamanid, H-isoniazid, hH-high dose isoniazid, stdH-standard dose isoniazid, LFX-levofloxacin, P-rifapentine, R-rifampicin
\*All TPT regimen formulations available under Global Drug Facility (GDF) procurement. Where a formulation is not listed under National Procurement, it is not available in South Africa under the national procurement mechanism (the Master Health Product List (MHPL))

There is substantial variation in cost between TPT regimens and between age and weight cohorts. Levofloxacin TPT dispensing solid 250 mg non-dispersible dosage tablets have a comparable cost to drug-susceptible TPT regimens across age ranges. Under GDF pricing, a TPT course using the 100 mg dispersible tablet formulation of levofloxacin is approximately 8-fold the price of the 250 mg tablet ($5 vs $38 in the 0-<5-year age range, and $79 vs $9 in the 5-<15-year age range). The levofloxacin dispersible tablet formulation is currently not yet purchased under South African national procurement, and the principal formulation of interest as used in the TB-CHAMP trial is the levofloxacin 250 mg solid dosage tablet. However, the optimised regimen reflected in Table 3 modelled the cost if the South African national programme obtained the levofloxacin 100mg dispersible tablet at GDF pricing. This demonstrated that if South Arica was able to access GDF supply of dispersible levofloxacin formulations only, and restricted use to lower age ranges, there would only be a marginal increase in costs compared to all children using the solid dosage levofloxacin formulation.

Potential future TPT regimens of delamanid and bedaquiline, if proven to be efficacious and safe, have substantially higher cost than levofloxacin, particularly under South African national pricing and for dispersible tablet formulations.

Total societal costs of TPT provision, incorporating pharmaceutical costs, other health system costs, and household costs for the 0-<5-year age range are shown in Figure 3 (with tabulated data and other age ranges in supplementary appendix). Pharmaceutical costs represent the largest variation between options, with non-pharmaceutical health system costs relatively stable between options (US$165 for clinical monitoring only to US$200 for 6-month regimen with monitoring). Non-pharmaceutical health system costs are predominantly driven by clinic visits, differentiated by visits associated with dispensing or monitoring, with regimen length determining overall costs. The analysis does not incorporate costs of commercial Interferon-Gamma Release Assays (IGRA) (US$110) or tuberculin skin testing (US$38)(29) as part of the pathway, as these are not required for initiation of TPT in current South African guidance (26); and were not required in the TB CHAMP protocol for the <5 age group. As a sensitivity analysis, the total health system costs for the <5 age groups increase to $307 and $236 depending on whether an IGRA or TST is used and increase in health system costs by 55% and 19% respectively.

**Figure 3.**
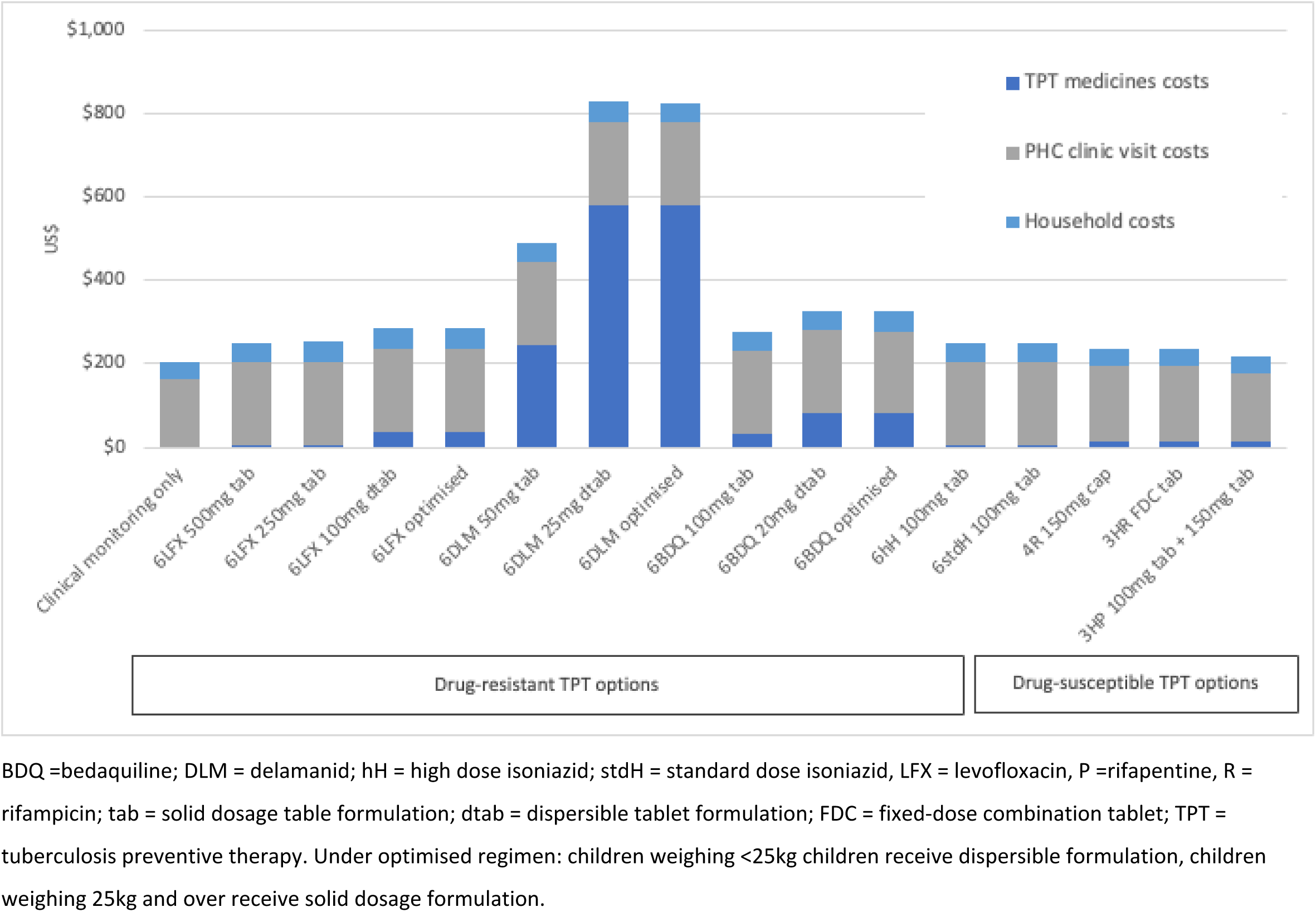
Total mean societal costs (US$) by TPT regimen and formulation for children 0-<5 years of age.

Household costs ranged from US$37-$48 across the whole treatment and monitoring period and were a small proportion of overall costs. Household costs were largely driven by duration of TPT regimen. Households participating in the TB-CHAMP trial reported a median total monthly income of US$186 in 2025 values (South African Rand (ZAR) 3,420), indicating that TPT costs represented 1.6%-2.1% of annual income in affected households over the treatment and monitoring period.

## DISCUSSION

This analysis details the cost of provision of a TPT strategy for children <15 years old exposed to MDR-TB in the South African context from a health system and household perspective. We demonstrate that factors including regimen choice, medicine formulation type, weight of the child, and procurement mechanism have a significant impact on the per patient pharmaceutical costs. The analysis combines empirical trial data for key parameters including pharmaceutical usage, clinic visits and weight distribution of participants, with normative recommendations on treatment regimens for medicines not included in the TB-CHAMP trial.

This analysis provides further context to existing work. A global modelling study by Dodd and colleagues assessed the cost effectiveness of TPT in household contact management in children and included regimen options that are represented in this analysis (34). Dodd and colleagues used cross country costs estimates based on GDF pricing for pharmaceuticals and per-country costs from WHO and generalised estimates based on GDP modifiers. An analysis by Jo and colleagues (35) assessed the costs and cost effectiveness of scale up of short-course TPT in child contacts of people with drug-susceptible TB across 12 countries, which also used global estimates modified across jurisdictions and secondary data from the Global Health Cost Consortium. Our analysis broadly aligns with the estimates from these studies, but provides further detail regarding the impact of different formulations, regimens, and weight bands on costs. We provide contemporaneous localised health system costs, and use empirical information on utilisation and adherence rather than rely solely on normative assumptions. This analysis also provides current pharmaceutical and other costing inputs for the year 2025; there has been substantial reduction in the costs of key antituberculosis agents (particularly bedaquiline and delamanid) in the previous three-year period.

Non-pharmaceutical health system costs predominantly comprised of clinic visits for monitoring and dispensing and constituted a substantial proportion of the costs, but were largely unaffected by regimen choice. Clearly shorter regimens and less intensive monitoring would reduce costs. Additional investigations at screening would substantially increase costs. A single IGRA test may provide useful additional information but would increase the health system costs per patient by 60%, far exceeding the cost of pharmaceutical treatment. This additional cost and further constraints on providing IGRA testing at scale supports the WHO guidance to initiate TPT for at-risk children below 5 years of age without the requirement for testing by TST or IGRA (26).

The household costs of providing TPT constitute the time to access clinics for monitoring and prescription pickup. These are relatively small, but may be a significant burden to households that may already be financially vulnerable. Importantly, household costs are not met by the national TB programme budget, but by general household expenditure. The MDRTBkids household survey established that for many households, a family member is able to walk to the local primary care clinic, avoiding direct travel costs. However, travel time and waiting time at clinics still represent a substantial opportunity cost for households.

Weight variation has a significant impact on per patient costs due to weight-based dosing, with differences in costs substantially higher for patients in the <5kg weight bracket (generally less than 2 months old) compared to children in the 20-25kg weight bracket (approximately 4 or 5 years old), with some variation by type of formulation. The inclusion of “optimised” regimens in the analysis attempted to reflect a practical scenario where children in the lower weight ranges (<25 kg) are provided dispersible (lower mg dose of medicine per unit) formulations, and those above are provided with adult formulations, would reduce the average cost of TPT across the age cohort. While this analysis reflects TB CHAMP weight-based dosing for levofloxacin, the recent harmonization of weight banding across tuberculosis treatment and prevention by WHO will enable further consolidation of dose schedules (23).

The analysis demonstrates that for levofloxacin, the use of a child-friendly formulation (100 mg dispersible tablet) is likely to be more expensive than the adult formulation (250 mg and 500mg solid tablet). While this is a high proportional difference and may be significant to the national TB programme at scale, the child-friendly formulation is expected to offer substantial additional benefits not captured in this analysis, including palatability, ease of administration, potential improved adherence (21,22) and accuracy of dosing given that the dispersible formulation would not require caregivers to halve or crush tablets. If South Africa were to procure (or receive through donor support) the 100 mg levofloxacin formulation, the unit price would be an important consideration in the sustainability of a national programme for TPT in MDR-TB exposed children.

The differences in prices secured through the national South African tender and the GDF are an important driver of pharmaceutical costs. The GDF procures the levofloxacin 250mg tablet at 28% of the price of the South African national procurement process, and across all formulations assessed in this analysis, GDF pricing was 70% of that achieved through South African procurement prices. It is expected that over time, the price of both delamanid and bedaquiline will reduce further following entry of generic manufacturers and ongoing competitive processes. Any policy decisions by South Africa’s national TB programme and Essential Drugs Programme related to use of levofloxacin or other regimen for TPT or MDR-TB disease treatment should take pricing and procurement dynamics into account, and particular consideration should be given to South Africa accessing GDF supply, particularly the 100mg dispersible tablet.

This analysis is purposefully South African based, drawing on empirical data from the TB-CHAMP trial, the MDRTBkids local household survey, and localised South African prices. The analysis models GDF pricing in addition to South African national tender pricing to facilitate transferability of the results, in particular the pharmaceutical costs to other national TB programmes that access GDF pricing. The transfer of total costs, however, would require modification of localised costs of screening, dispensing and primary health clinic monitoring visits.

The analysis has some limitations. Most costs are based on empirical trial or current real prices (such as the South African tender and national laboratory price list); however, costs based on non-empirical data such as costs of primary care clinic visits required secondary data and inflation of reported costs to reflect 2025 prices. As with all costing analysis, prices, and particularly pharmaceutical prices, are subject to change associated with procurement cycles, and should be interpreted with reference to dates of cited pricing sources. In line with established practice, costs specific to the operational aspects of the TB CHAMP were excluded from the costs as these would not be replicated at implementation bit it is possible that some aspects of the clinical trial influenced outcomes such as adherence. This should be taken into consideration in any secondary analysis utilising costs parameters in this analysis.

Our analysis reflects current MDR-TB TPT clinical guidance with potential future regimens for comparison. Costs may significantly decrease with the introduction of shorter regimens, options with fewer follow-up or monitoring visits, and further competition in the market for key proprietary pharmaceuticals.

The analysis is also broadly representative of what would be expected in a real-world scaled-up scenario. The empirical data is from a clinical trial, so may overestimate adherence to treatment over the 6-month period or feasibility of the health system to provide monitoring over the 2-year period. However, the data may be more reflective of actual implementation compared to a completely normative ingredients-based costing analysis that implicitly assumes 100% compliance with recommended guidance. This analysis also did not represent cost efficiencies that may be achievable with integration of TPT with other programmes to dispense medication or monitor patients (such as antiretroviral treatment). It is expected that it would be possible for household costs to be reduced if a family member was able to collect medication for both the member receiving treatment for MDR-TB and the child’s TPT at the same visit.

Our analysis excludes some cost elements that may be relevant to some users. The “above service delivery” costs, such as the managerial costs of organising the TPT programme have not been included but may reasonably be considered to fall within existing functions of the TB programme and health managers. Importantly, the “above service delivery” costs are not expected to vary significantly between different TPT options. Contact tracing to identify children for TPT initiation can have substantial human resource costs including community health workers, administrators and clinicians and should be incorporated if assessing a wider prevention programme. Medicine administration of adult formulations (such as a solid tablet) for children in this age group commonly involves crushing the tablet and suspending in water, yoghurt, or other palatable media; administration of dispersible formulation involves suspending the tablet in water. The health professional or caregiver costs associated with administration of the different formulations was not included in the analysis due to uncertainty associated with estimation and attribution of costs; however, it is likely that the convenience of the dispersible formulations represent slightly lower administration cost. As the TB-CHAMP trial which included almost 1000 children did not demonstrate a significant difference in adverse events between levofloxacin and placebo, the cost estimates in this analysis do not incorporate costs to health systems or households of adverse event management. Secondary analysis or use of the costs should acknowledge this assumption and add costs of adverse event management if adverse events of TPT are found to be significant in future.

In conclusion, this is the first analysis that utilises empirical data and localised costs and prices to estimate the expected per patient costs of TPT for MDR-TB in South African children, across a range of potential treatment options. The results will contribute to existing initiatives to collate and disseminate TB cost parameters (17), and the findings will be integral inputs to budget planning and further estimates of the cost effectiveness of different strategies and feasibility of implementation in the South African and other settings.

## Data Availability

The data supporting this costing synthesis analysis is all in public domain in cited price schedules or cited publications.

## Notes

### Competing Interest Statement

The authors have declared no competing interest.

### Funding Statement

Yes

### Author Declarations

Stellenbosch University Health Research Ethics Committee

